# Disentangling the distal association between β-Amyloid and tau pathology at varying stages of tau deposition

**DOI:** 10.1101/2023.03.31.23288013

**Authors:** Seyed Hani Hojjati, Gloria C. Chiang, Tracy A. Butler, Mony De Leon, Ajay Gupta, Yi Li, Mert R. Sabuncu, Farnia Feiz, Siddharth Nayak, Jacob Shteingart, Sindy Ozoria, Saman Gholipour Picha, Antonio Fernández, Yaakov Stern, José A. Luchsinger, Davangere P. Devanand, Qolamreza R. Razlighi

## Abstract

Studies on the histopathology of Alzheimer’s disease (AD) strongly suggest that extracellular β-amyloid (Aβ) plaques promote the spread of neurofibrillary tau tangles. Despite well-documented spatial discrepancies between these two proteinopathies, their association remains elusive. In this study, we aimed to investigate the distal (non-local) association between tau and Aβ deposition by studying the Aβ, and tau positron emission tomography (PET) scans of 572 elderly subjects, aged 67.11 ± 6.08 years (476 healthy controls (HC), 14 with mild cognitive impairment (MCI), 82 mild AD). We also leveraged 47 tau-PET and 97 Aβ-PET scans of healthy young individuals (aged 20-40) to find regional cut-points for tau- and Aβ-positivity in 68 cortical regions in the brain. Based on these cut-points, we implemented a pseudo longitudinal technique to categorize the elderly subjects into four pathologic phases of AD progression: a no-tau phase, a pre-acceleration phase, an acceleration phase, and a post-acceleration phase. We then assessed the distal association between tau and Aβ in each phase using multiple linear regression models. First, we show that the association between tau and Aβ starts distally in medial temporal lobe (MTL) regions of tau (e.g., left and right entorhinal cortex and right parahippocampal gyrus) in the early stage of tau aggregation (pre-acceleration phase). We then show that tau in several bilateral brain regions (particularly the entorhinal cortex and parahippocampal gyrus) exhibits strong distal associations with Aβ in several cortical brain regions during the acceleration phase. We found a weak distal association in the post-acceleration phase, comprising 96% of MCI or mild AD and Aβ+ subjects. Most importantly, we show that the HC Aβ+ subjects have the highest degree of distal association between tau and Aβ of all the subjects in the acceleration phase. The results of this study characterize the distal association between the two key proteinopathies of AD. This information has potential use for understanding disease progression in the brain and for the development of anti-tau therapeutic agents.

## Introduction

The pathology of Alzheimer’s disease (AD) is characterized by two key proteinopathies: intracellular neurofibrillary tau tangles and extracellular β-amyloid (Aβ) plaques [1, 2]. Both of these proteins accumulate and spread with time as the disease progresses. It has been found that these proteins aggregate in both cognitively normal elderly and AD patients [3]; also, the trajectory of tau accumulation differs significantly between AD patients [4]. The advent of the second generation of positron emission tomography (PET) tau and Aβ tracers enables us to accurately investigate the associations between tau and Aβ deposition from the cognitively healthy to AD dementia stages of the disease to characterize how these proteinopathies interact during AD pathogenesis.

Postmortem data indicate that tau deposition tends to have a region-specific progression that initiates in the entorhinal cortex (specifically in the transentorhinal cortex), where negligible Aβ pathology is found [4]. Six consecutive stages of tau deposition have been described to occur over the stages of AD progression (Braak staging regions) [5-7]. Individuals in Braak stages I-II typically have no clinical symptoms, with tau proteins restricted in the medial temporal lobe, and cognitive deficits typically emerge from stage III [5-7]. However, neither PET studies nor autopsy data have identified the region-specific progression from which Aβ deposition initiates and spreads during the progression from normal aging to AD. Aβ is shown to accumulate ubiquitously throughout the cingulate, medial parietal, and prefrontal cortices, where negligible tau pathology is found in the early stages of tau accumulation [8, 9].

The mechanisms that cause tau to spread out of the transentorhinal and entorhinal cortex and later spread out of temporal and limbic regions are key unsolved questions. Although tau tangles and Aβ plaques are considered the primary indicators of AD pathology, there is only a weak correlation between Aβ and cognition [10]. In contrast, tau strongly predicts cognitive decline [11-13], and neuropathological data suggest that tau triggers the relationship between Aβ and cognitive decline in MCI and AD patients [14, 15]. Furthermore, the early stages of tau deposition in transentorhinal cortex are commonly observed in older healthy subjects (aged>60), while the spread of tau to limbic and neocortical areas is usually complemented by the presence of Aβ plaques [16, 17]. Previous studies also propose two stages of tau deposition: the age-related stage and the pathological stage. The accumulation of tau pathology as a result of aging is limited to the medial temporal lobe and does not spread to the isocortical Braak stages [18]. On the other hand, the pathological stage of tau is not restricted to the medial temporal lobe (MTL) region. Notably, early tau deposition is restricted to the MTL and limbic regions without Aβ pathology [18]. Most of the previous studies as well as our work [19], have considered the local association between Aβ and tau pathologies in the stages in which these two proteins spatially overlap in the brain. However, spatial discrepancies between the early deposition of these two proteins and the distal association between these two proteins remain less understood when Aβ and tau are not spatially overlapped in the brain. In addition, it remains unclear whether Aβ and tau have distal interactive effects, can accelerate the spreading of pathology, and lead to more cognitive decline.

The main aim of this study was to characterize the distal association between Aβ and tau in different stages of tau accumulation. Notably, we want to study intra-regional associations among regions’ tau and Aβ accumulation in various stages. We hypothesize that the association between tau and Aβ starts distally in non-local connected regions in the early stage of tau accumulation.

## Method

### Participants

Five hundred and seventy-two older (aged > 55 years) and 144 younger (aged 20-39 years) subjects from eight research cohorts from the Weill Cornell Medicine and Columbia University Irving Medical Center were included in this study. The older cohort included 477 HC, 14 MCI, 82 mild AD participants (342 females) who underwent three different imaging scans (3T T1-weighted structural MRI, tau PET, and Aβ PET) within 12 months. Younger subjects had either a tau PET (47 subjects) or Aβ PET (97 subjects) scan, as well as a 3T T1-weighted structural MRI scan. These young subjects were used as a normative reference group to generate the region-specific cut-points for each protein. All participants gave informed consent to participate in their respective studies, and the local institutional review boards approved all recruitment/enrollment procedures and imaging protocols. The younger and older HC subjects underwent medical and neuropsychological evaluations to confirm the don’t have any neurological or psychiatric conditions, cognitive impairments, major medical illnesses, or any contraindications based on structural images. The patients with MCI or mild AD had Mini-Mental State examination (MMSE) scores ranging between 18 and 28, a Clinical Dementia Rating (CDR) of 0.5 (MCI) or 1.0 (mild AD), and the presence of a biomarker associated with AD (either a positive Aβ PET scan or cerebrospinal fluid (CSF) analysis showing positivity for Aβ42, tau, and phospho-tau protein181.

### MR and PET image acquisition protocols

For tau-PET, all subjects were injected with 185 MBq (5 mCi) ± 20% (maximum volume 10 mL) of 18F-MK6240 before imaging that was administered as a slow single IV bolus at 60 seconds or less (6 secs/mL max. Imaging was performed as six 5-min frames for a 30-min PET acquisition, 80–120 min post-injection. If it was considered inadequate, an additional 20 minutes of continuous imaging were performed.

For Aβ imaging, subjects underwent 18F-Florbetaben or 18F-Florbetapir PET scans. This scan consisted of four 5-min frames over 20 minutes of acquisition, starting 50–90 min after injection of 8.1mCi ± 20% (300 MBq) of the tracer, which was administered as a slow single IV bolus at 60 seconds or less (6 secs/mL max).

MRI scans at 3T with a 3D volumetric T1 magnetization-prepared rapid gradient-echo sequence was performed. Each subject first underwent a scout localizer to determine the position and set the field of view and orientation, followed by a high-resolution MPRAGE image with TR /TE = 2300-3000/2.96-6.5 ms, flip angle = 8-9°; field of view = 25.4-26 cm, and 165-208 slices with 1 mm thickness.

### Neuroimaging preprocessing

All MPRAGE scans were processed with FreeSurfer 7.1.0 (http://surfer.nmr.mgh.harvard.edu) for automated segmentation and cortical parcellation (e.g., including segmentation, and creation of an average gray matter mask) [20, 21] to derive regions of interest (ROIs) in each subject’s native space using the Desikan–Killiany atlas [22]. These ROIs were utilized for calculating Aβ and tau PET regional measures. The Aβ and tau regional standardized uptake value ratio (SUVR) were calculated by normalization to cerebellum grey matter. ROIs of this study are based on 68 cortical regions of the Desikan-Killiany atlas.

The fully automated in-house pipelines were utilized to process the Aβ and tau PET images as previously published [23-27]. Both Aβ and tau PET dynamic frames (six frames in tau PET and four in Aβ PET) were aligned to the first frame using rigid-body registration and averaged to generate a static PET image. Next, the structural T1 image in FreeSurfer space was registered to the same subject’s PET composite image using normalized mutual information and six degrees of freedom to obtain a rigid-body transformation matrix.

We have developed a method to correct partial volume effects in Aβ PET scans using a normative reference group of young individuals aged 20 to 40 years. Our technique uses the gray matter voxel uptake to estimate the off-target binding from white matter for florbetaben scans. While we do not expect to see cortical Aβ in this young reference group, we can use their data to model the off-target binding in gray matter. We extracted the white matter uptake from test subjects and used it to estimate the synthesized spill-in for the gray-matter voxels, which was then subtracted from the actual uptake to reduce artifacts from white matter regions. This process was done using fitted model parameters at each voxel. Overall, our anatomy-driven partial volume correction technique effectively reduces the impact of white matter on Aβ PET scans.

To classify the Aβ+ subjects, we generated the global cortical Aβ, including frontal, parietal, temporal, anterior cingulate, posterior cingulate, and precuneus ROIs [28-30]. Next, we calculated the normal distribution of global Aβ values in the younger participants. Finally, using the 95^th^ percentile of the fitted normal distribution, we calculated the cut-point for defining abnormal global Aβ (cut-point=1.2508). By generating the same regions of interest for older subjects and utilizing the cut-point=1.2508, we classified the Aβ+ (global Aβ >1.2508) subjects.

### Regional cut-points for Aβ and Tau

Most existing studies utilize global cut-points to separate negative and positive subjects based on their global Aβ and tau pathologies [31]. Using this method has led to varying cut-points in different studies and completely disregards regional elevation of Aβ and tau, which can carry critical information on the regional evolution of the disease, particularly in HC subjects. In addition, off-target binding of Mk-6240 tau tracer in meningeal has a significant impact on the cortical SUVR data [32]. By considering the global cut-points, it is difficult to account for off-target binding, which may differ from region to region in the cortex. In this study, instead of utilizing the conventional global cut-points, we developed a technique to calculate regional cut-points for each cortical region of Aβ and tau deposition relative to deposition in the normative reference group. We used this regional cut-point to define the different stages of tau accumulation and categorize the subjects by considering the regional positivity and spatial distribution of tau protein. Notably, we only use the Aβ regional cut-points to visualize Aβ spatial distribution in different groups of subjects. To determine the regional cut-points using the selected atlas (Desikan Killiany) with 68 cortical regions, we calculated the normal distributions of the normative reference group regional tau and Aβ SUVR. Then for each region, we determined the 95th percentile of the fitted normal distribution as regional cut-points. The regional cut-points for tau based on the Desikan Killiany atlas and normative reference group vary between about 0.9 to 1.3. Thus, the regions closer to the meningeal off-target binding have higher cut-point values, and the regions far from the meningeal of target binding will have lower cut-point values (Figure 1).

**Figure 1.**
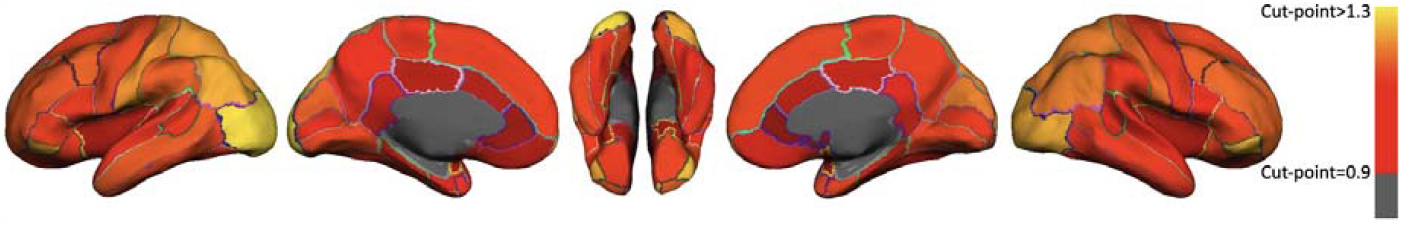
Illustrating the regional cut points of tau pathology throughout the 68 cortical regions. The cut-points for each region are displayed using a heat color map and overlaid on the semi-inflated cortical surface of the MNI152 template for visual representation. The red color indicates a cut-point value equal to 0.9, and the yellow color indicates a cut-point value higher than 1.3 for each cortical region.

### Pseudo longitudinal categorization of the elderly subjects

For categorizing the older subjects, first, we used the obtained regional tau cut-points and determined the number of regions in each subject that exceeded the regional cut-point. Then we ordered the subjects based on the number of regions that exceeded the regional cut-points (from 0 to 68).

Regions were also sorted by the frequency at which the regional cut-point was crossed across all subjects to define a regional tau spreading order (Figure 2). We defined three different thresholds to categorize the subjects into four phases: no tau, pre-acceleration, acceleration, and post-acceleration. The “no-tau” phase includes subjects with no regions exceeding the regional cut-points. Once a subject exceeds a single region cut-point, the first threshold is considered crossed, separating the no-tau phase and pre-acceleration phase subjects. We utilized the elbow method for the second threshold, at which the smoothed graph showed the highest change (pre-acceleration phase). Finally, the last threshold separated the two last phases of subjects, where the second derivative became zero. This threshold separated the acceleration phase and post-acceleration phase, see Figure 2.

**Figure 2.**
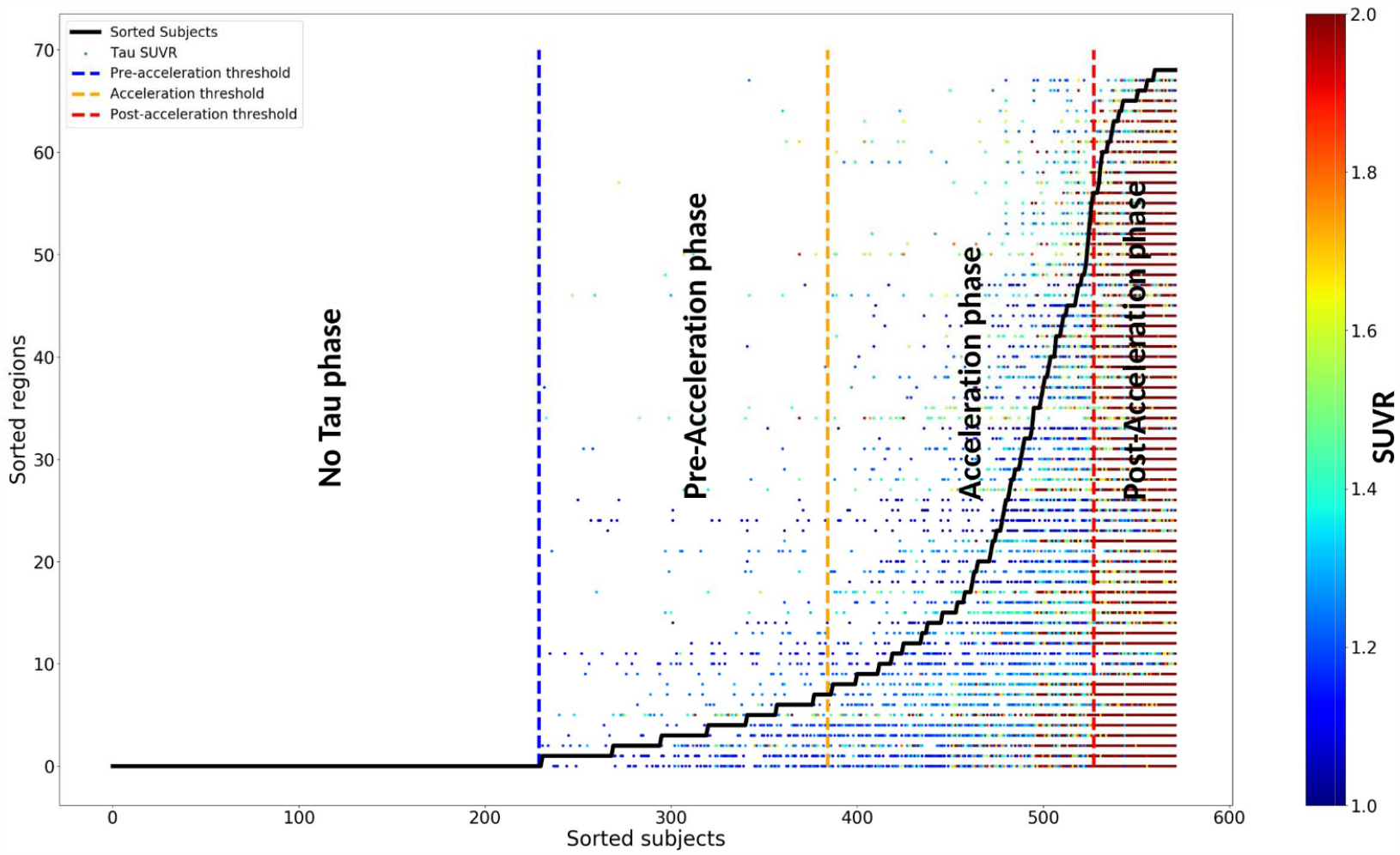
The sequence of tau deposition in 572 elderly subjects based on young subjects’ regional cut-point. Subjects were sorted based on the number of tau-PET regions that exceed the regional cut-points in the x-axis. Regions were also sorted by the frequencies exceeding the regional cut-point across all subjects in the y-axis. The tau SUVR for the regions that exceed the regional cut-point is color-coded with a heat map; the blue color indicates the tau SUVR value is equal to 1, and the red color indicates the tau SUVR values higher than 2. Three thresholds were defined to separate the subjects into regions into four phases no-tau, pre-acceleration, acceleration, and post-acceleration.

### Statistical analysis

After categorizing the older subjects, a probabilistic atlas was obtained to visualize the Aβ and tau deposition pattern in each category. For visualization, we applied the regional cut-points of Aβ and tau and then binarized each region. Lastly, we calculated the probability of observing abnormal Aβ/tau levels (as determined by cut-off points) among individuals in each category.

The distal association Aβ and tau depositions were assessed in all 68 target regions. We applied a multiple regression model for each target region of tau (i) to assess the association with Aβ deposition in all other regions (j) (67 independent multiple regression analyses, *i* ≠ *j*), while age, gender, ICV, and target region Aβ were controlled as covariates:

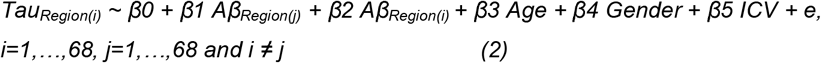

Finally, for each group and target region, statistical maps (t-test) were generated to visualize regions with significant distal associations between the target region of tau and the other regions’ Aβ depositions.

This study used Python for all statistical analyses and visualizations. The main numeric modules and visualization were utilized, NumPy and Matplotlib [33, 34]. Statistical tests such as analysis of variance (ANOVA) and Chi-square tests were performed using the SciPy statistical package (v6.1.1) [35]. A permutation test performed family-wise error correction of regional associations. A null distribution was determined by randomly shuffling the independent variable 10,000 times. Based on the 95th percentiles of the fitted normal distribution for positive t-values, we calculated the family-wise error rate-corrected t-value.

## Results

### Subject characteristics and categorization

The regional distributions of tau and Aβ in elderly subjects reflecting the four phases of tau deposition (no tau, pre-acceleration, acceleration, and post-acceleration) are depicted in Figure 3. Table 1 illustrates the number of subjects and their demographic characteristics in each phase. The regional distributions of tau and Aβ in different phases show interesting topographical differences. As shown in Figure 3 and Table 1, the no-tau phase subjects comprised only about 4% of subjects with MCI or mild AD and 18% with a limited amount of Aβ deposition (frontal and temporal lobes). However, based on Figure 2, in the pre-acceleration phase, tau deposition existed in 8 regions and was largely restricted to certain MTL sub-regions (e.g., entorhinal cortex, parahippocampal gyrus); this group was comprised of 7% MCI or mild AD and 19% Aβ+ subjects. Aβ and tau deposition and spatial distribution increased in the acceleration phase, and except for ten cortical regions, all other regions showed a significant amount of tau in this phase. Subjects in this phase comprised about 22% MCI or mild AD and 45% Aβ+ subjects. Finally, tau and Aβ dominated whole cortical regions in the last phase (post-acceleration phase); about 96% of subjects in this group are MCI or mild AD, and Aβ+. Throughout different phases of tau deposition, Aβ and tau levels of accumulation, distribution, and disease stages gradually worsen from the first to the last phase.

**Table 1.** cohort demographics

| | Young Tau PET Participants (N=47) | Young Aβ PET Participants (N=97) | Old Aβ/Tau PET participants (N=572) | No Tau (N=229) | Pre-Acceleration Phase (N=155) | Acceleration Phase (N=143) | Post-Acceleration Phase (N=45) | Four group difference (P-value) |
| --- | --- | --- | --- | --- | --- | --- | --- | --- |
| Age (years) | 29.36 ± 4.73 | 27.64 ± 3.23 | 67.11 ± 6.08 | 65.52 ± 4.72 | 67.12 ± 5.78 | 69.27 ± 6.99 | 68.24 ± 7.60 | P<0.0001 |
| Sex (M/F) | 20/27 | 42/55 | 230/342 | 101/128 | 61/94 | 50/93 | 18/27 | P=0.372 |
| HC/MCI/Mild AD | 47/0/0 | 97/0/0 | 476/14/82 | 220/5/5 | 143/2/9 | 111/3/29 | 2/4/39 | P<0.0001 |
| Aβ positive/Aβ negative | - | - | 177/395 | 41/188 | 29/126 | 64/79 | 43/2 | P<0.0001 |

Abbreviations: HC: healthy control, MCI: mild cognitive impairment, M: male, F: female, PET: positron emission tomography, Aβ : β-amyloid

**Figure 3.**
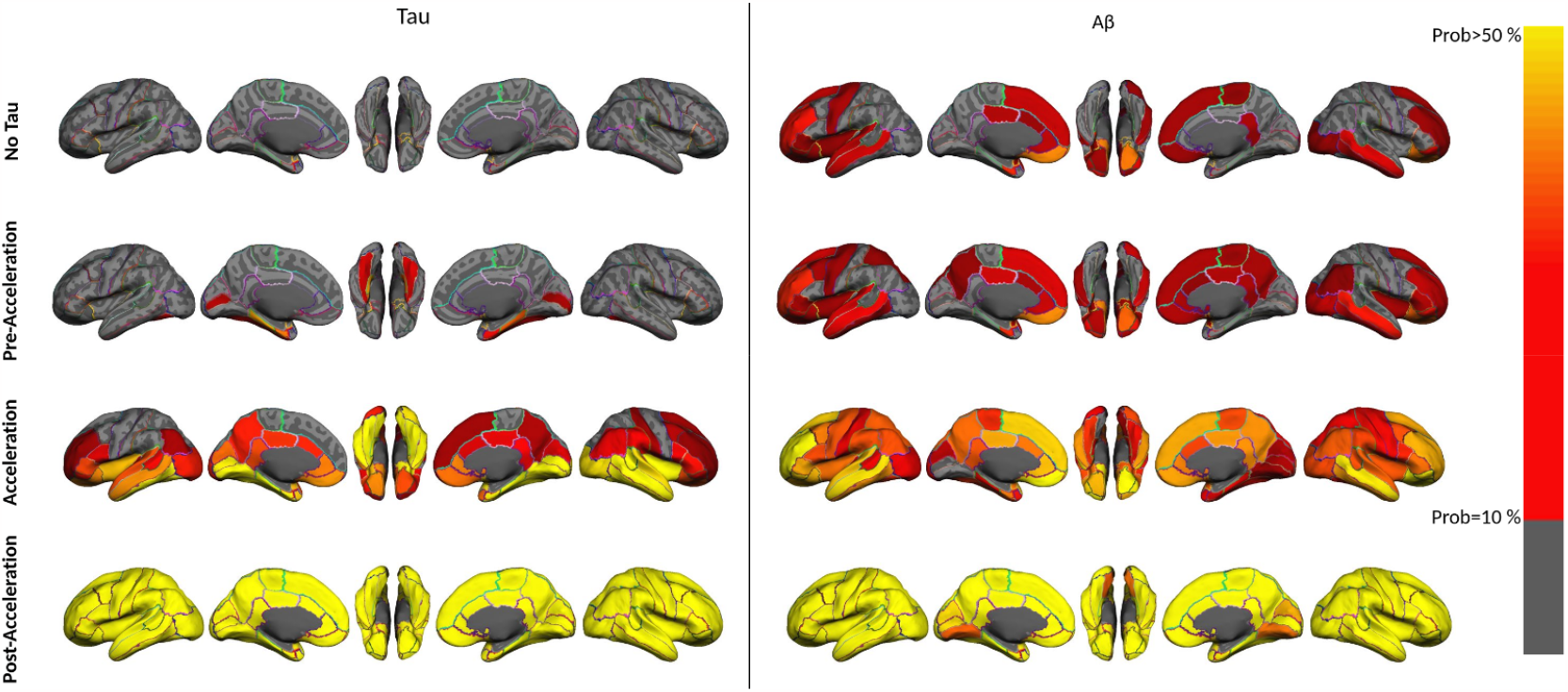
Illustrating the region-wise probabilistic atlas of tau (left column) and Aβ (right column) pathologies throughout the cerebral cortex obtained in four phases of tau accumulation. (First row, no-tau; second row, pre-acceleration; third row, acceleration; and fourth row, post-acceleration). The probability of observing tau and Aβ at each region is displayed using a heat color map and overlaid on the semi-inflated cortical surface of the MNI152 template for visual representation. The red color indicates a probability value equal to 10% of the subjects, and the yellow color indicates a probability value higher than 50%.

### Distal association in four phases of tau depositions

To explore whether distal Aβ deposition influenced tau deposition, we applied analyses on 68 target regions. As expected, no statistically significant association survived between Aβ and tau deposition in the no-tau phase. This is evident due to the absence of tau deposits in this phase after applying the regional cut-points. Distal association analysis captured three target regions in the pre-acceleration phase (Figure 4) that showed a significant distal association with several regions’ Aβ uptake, left and right of the entorhinal cortex, and right parahippocampal gyrus. The right entorhinal cortex is strongly associated with several Aβ regions, including the bilateral middle temporal gyrus, inferior temporal gyrus, and fusiform gyrus. This result can be implied as evidence of a distal association between early tau deposition in the MTL region and Aβ outside the MTL region.

**Figure 4.**
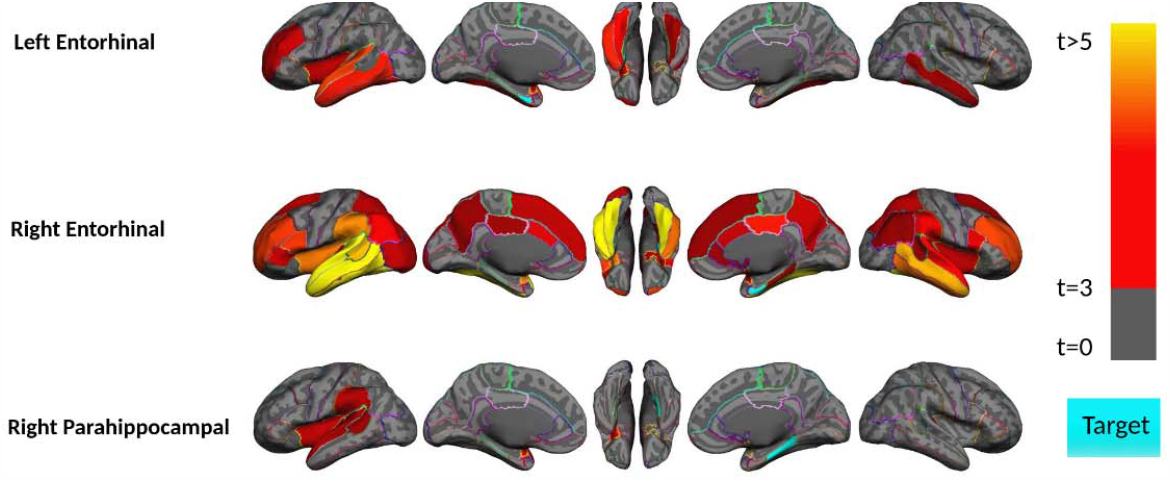
Region-wise statistical map (t-value) of distal association between tau deposition in three target regions and regional Aβ depositions in 67 cortical regions obtained in the pre-acceleration phase of tau deposition. The family-wise corrected t-value at each region is color-coded with red or yellow colors representing increasing positive t-values and overlaid on the semi-inflated cortical surface of the MNI152 template. The red color indicates the t-value equal to 3, and the yellow color indicates the t-values higher than 5. The target region indicates as light blue.

The acceleration phase subjects displayed the strongest distal association between Aβ and tau depositions among all phases of tau deposition (Figure 5). There was a significant association between 28 out of 68 tau target regions. However, the bilateral entorhinal cortex and parahippocampal gyrus appear to have the highest distal associations in the acceleration phase.

**Figure 5.**
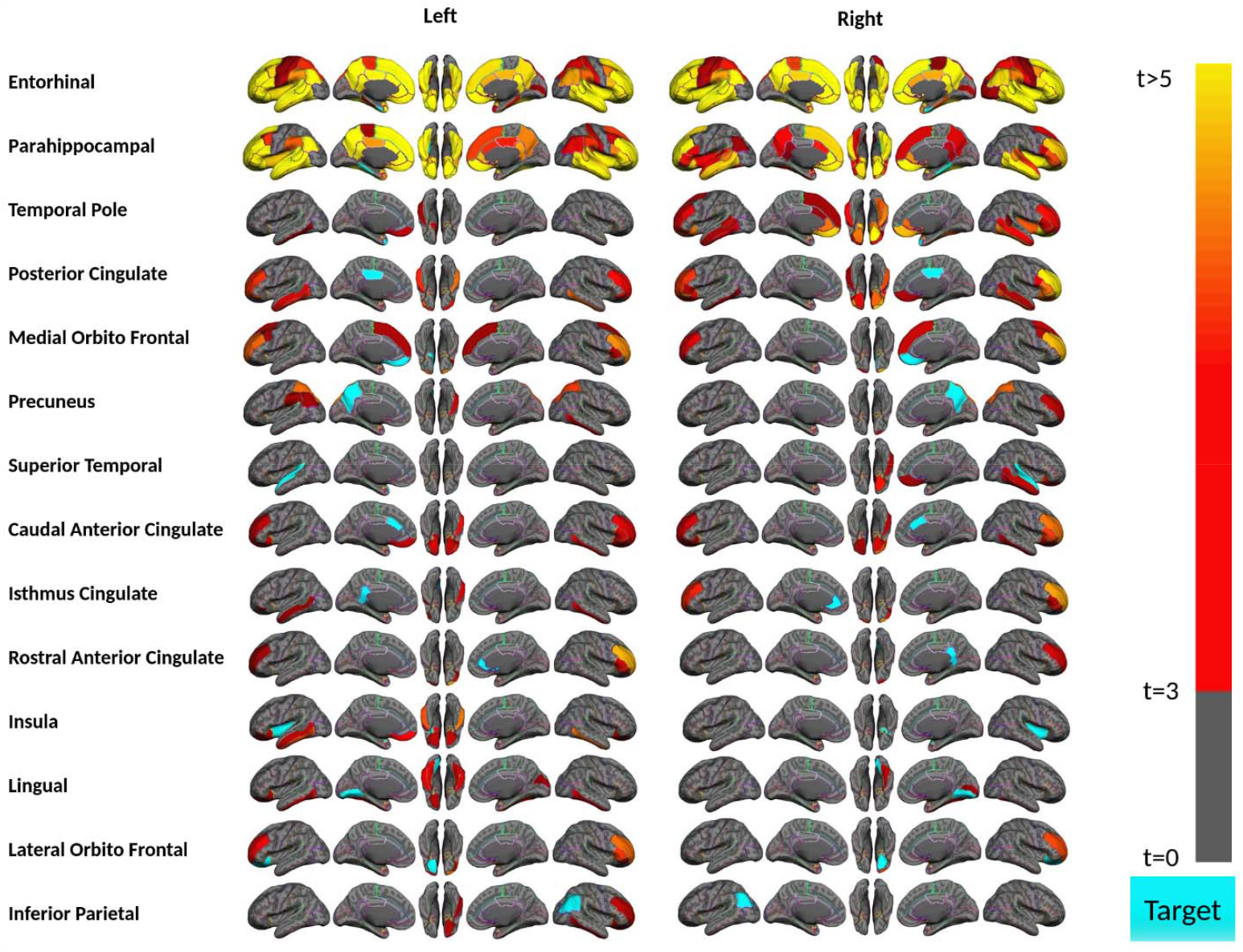
Region-wise statistical map (t-value) of distal association between tau deposition in fourteen target regions and regional Aβ depositions in 67 cortical regions obtained in the acceleration phase of tau deposition. The family-wise corrected t-value at each region is color-coded with red or yellow colors representing increasing positive t-values and overlaid on the semi-inflated cortical surface of the MNI152 template. The red color indicates the t-value equal to 3, and the yellow color indicates the t-values higher than 5. The target region indicates as light blue.

These two bilateral target regions illustrate the significant associations with more than 55 regions of Aβ deposition. Tau deposition in this phase is more likely to be related to Aβ deposition in other regions compared to the other three phases. Surprisingly, no significant distal association survived multiple comparisons in the post-acceleration phase; the tau and Aβ deposition were elevated in this phase.

### Aβ+ HC subjects show the highest distal association

Based on previous sections, the acceleration phase was characterized by a strong distal association. To investigate which subset of subjects in the acceleration phase has the strongest association, we conducted several additional analyses. We first separated the acceleration phase subjects into two HC (111 subjects with 37 Aβ+ subjects) and MCI or mild AD (32 subjects with 27 Aβ+ patients) groups. Subsequent analyses were conducted across MCI and mild AD subjects to determine how AD severity affected the association. Figure 6 shows this experiment results across MCI and mild AD subjects during the acceleration phase. Tau in five target regions illustrates weak associations with limited non-local Aβ brain regions.

**Figure 6.**
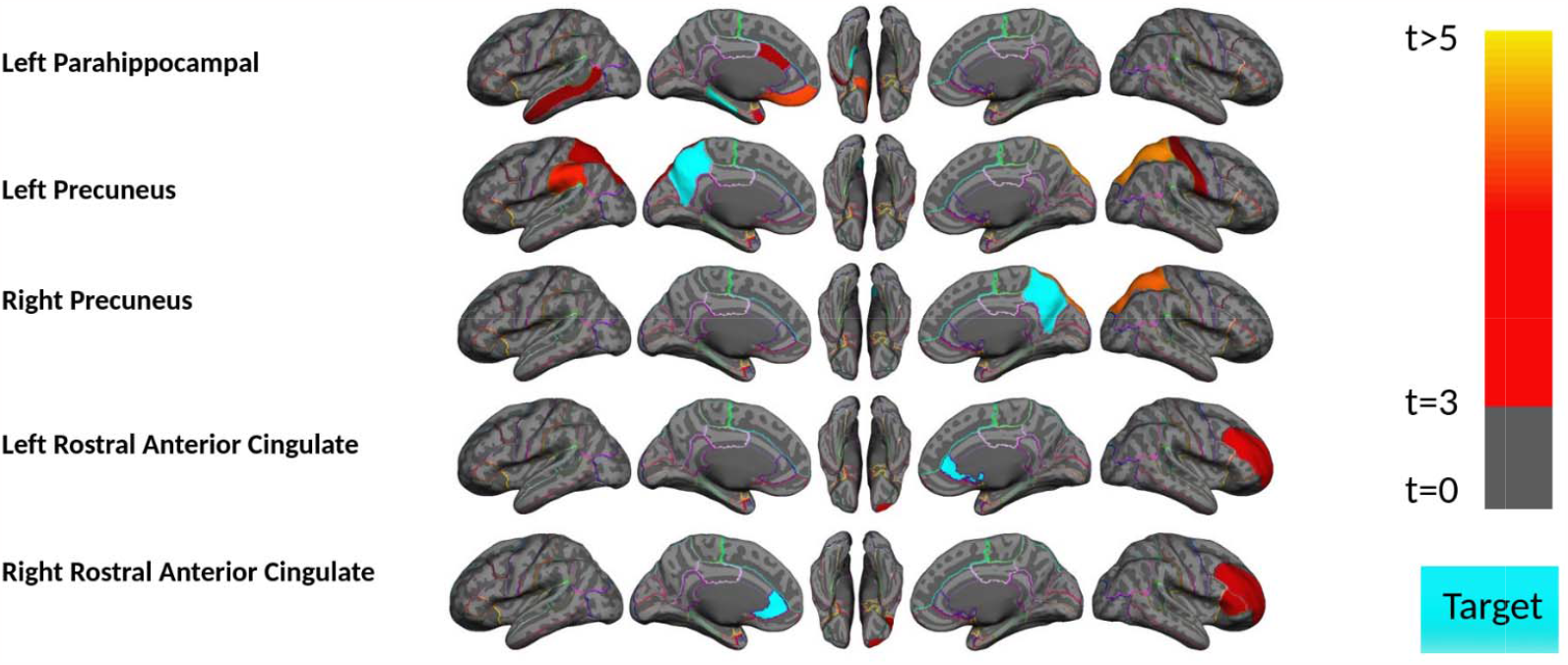
Region-wise statistical map (t-value) of distal association between tau deposition in five target regions and regional Aβ depositions in 67 cortical regions obtained the MCI and mild AD subject in acceleration phase of tau deposition. The family-wise corrected t-value at each region is color-coded with red or yellow colors representing increasing positive t-values and overlaid on the semi-inflated cortical surface of the MNI152 template. The red color indicates the t-value equal to 3, and the yellow color indicates the t-values higher than 5. The target region indicates as light blue.

For further investigation on HC subjects, we next assessed whether the Aβ deposition level alters the obtained distal association. Therefore, we divided the 111 HC acceleration subjects into 37 Aβ+ and 74 Aβ-groups and investigated the associations separately. In HC Aβ-subjects, only two target regions (left lingual and right entorhinal cortex) of tau show marginal associations with non-local Aβ (Figure 7). On the other hand, tau in six regions shows a significant association with Aβ in several brain regions in HC Aβ+ subjects. Interestingly, the right entorhinal cortex strongly associates with non-local Aβ in several regions. Once again, these results confirm that distal association becomes considerably weaker after the disease onset (impaired subjects), whereas at the preclinical stage, the existing Aβ deposition demonstrates a strong distal association with tau.

**Figure 7.**
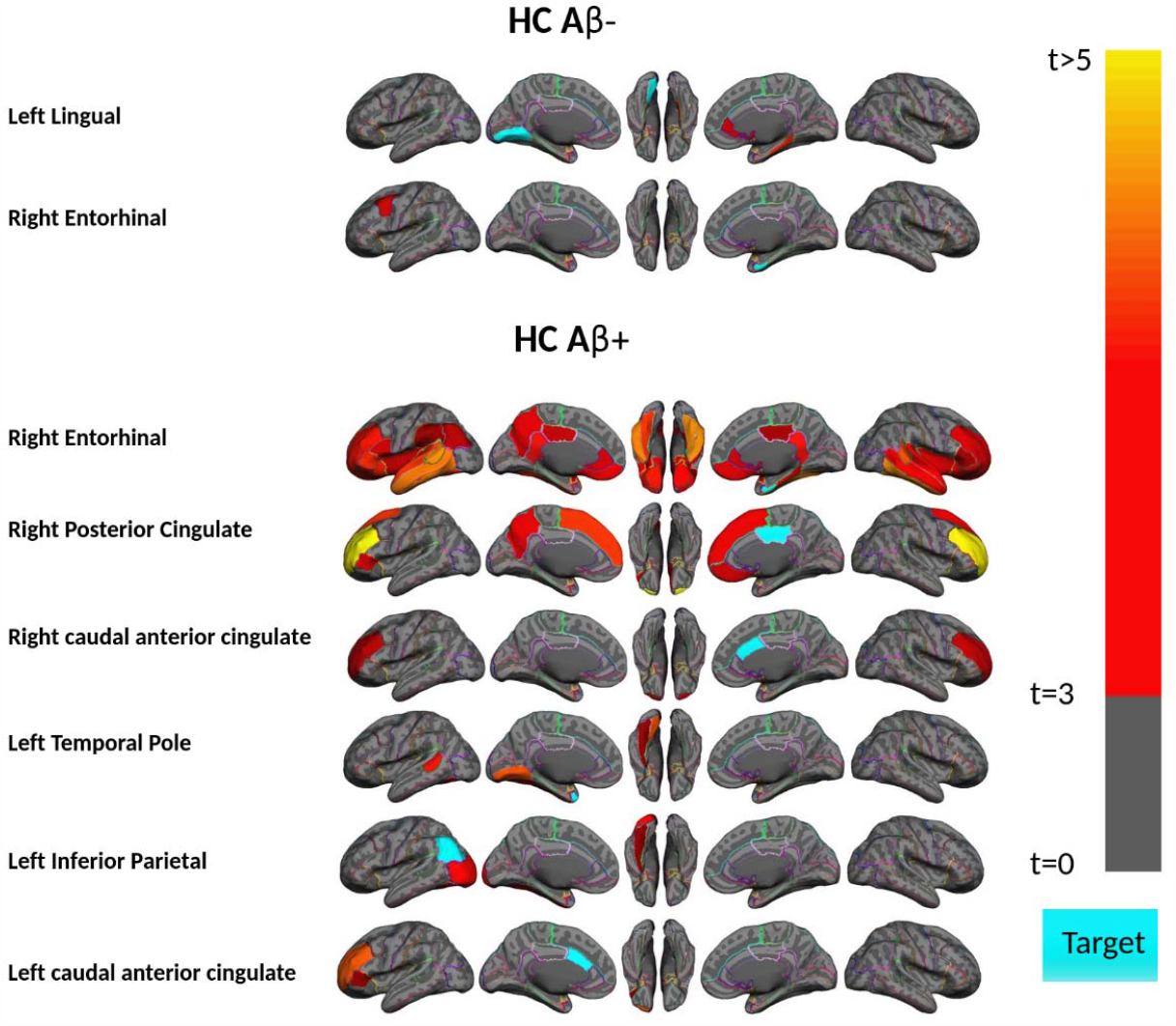
Region-wise statistical map (t-value) of distal association between tau deposition in six target regions and regional Aβ depositions in 67 cortical regions obtained in the HC Aβ- and Aβ+ subjects in the acceleration phase of tau deposition. The family-wise corrected t-value at each region is color-coded with red or yellow colors representing increasing positive t-values and overlaid on the semi-inflated cortical surface of the MNI152 template. The red color indicates the t-value equal to 3, and the yellow color indicates the t-values higher than 5. The target region indicates as light blue.

### Discussion

Since the accumulation of AD pathology is gradual and often starts decades before the onset of the disease, it is crucial to understand the progression of neuropathology during aging in order to prevent the development of AD and its clinical implications. In this study, we implement the pseudo longitudinal technique to assess the distal association between tau and Aβ deposition within 68 brain regions. The association was assessed throughout four phases of tau deposition based on our pseudo longitudinal categorization method: no-tau phase, pre-acceleration phase, acceleration phase, and post-acceleration phase. The main findings in this study are: First, the association between tau and Aβ starts distally in the early stages of tau deposition in the MTL region. The second finding is that this association accelerates and continues in the next stages of accumulation (acceleration phase), but then declines significantly in the last/symptomatic stage of the disease.

Third, tau deposition was strongly associated with the entorhinal cortex and parahippocampal gyrus, which appear as hub regions for the distal associations throughout the early to middle stages of tau deposition. Fourth, the results of HC Aβ+ subjects in the acceleration phase indicate that the strongest associations happen in the HC subjects with abnormal levels of Aβ deposition in the acceleration phase. Finally, surprisingly in the disease stage (MCI and mild AD subjects), the association between tau and Aβ deposition was weak.

Several studies have revealed that tau deposition begins in the entorhinal cortex during normal aging [36-39], and spreads throughout the MTL in the asymptomatic preclinical AD stages. The amyloid cascade hypothesis indicates that tau pathology can be found within MTL during normal aging in the absence of significant neocortical Aβ deposition, and Aβ pathology acts as a gatekeeper for tau pathology to spread out of the MTL to the neocortex [40]. Our results in the pre-acceleration phase suggest that the entorhinal cortex and parahippocampal gyrus tau have the highest distal associations even at this early deposition stage. The association between these two regions’ tau and Aβ would suggest that the association between these two pathologies happens earlier than the literature reported. The difference is that the association of tau inside of the MTL tau happens distally with Aβ, and Aβ may enhance the tau spreading even inside of MTL via distal association. The distal association of the entorhinal cortex tau with other cortical regions has been supported by previous studies that showed that tau spreads to regions functionally and structurally connected to the entorhinal cortex [41-44]. This association may explain the spreading of tau deposition following functional connections [45-47]. The distal association is also shown by in-vivo studies that indicated that tau spreads to distal but functionally connected brain regions, affecting intrinsic functional neuronal networks [46-48]. Thus, although age-related MTL tau accumulation is well-documented as a substrate of tau spreading, the distal association between tau and Aβ is critical to understand how tau spreads and progresses in the early stages of accumulation.

Consistent with the above findings, in the acceleration phase, strong distal associations appeared in several tau target regions. More importantly, the spreading of the tau requires the presence of abnormal Aβ (positivity), as we observed strongest associations across the HC Aβ+ subjects in acceleration phase. Previous reports also observed that tau increased faster with higher Aβ deposition than lower Aβ deposition in clinically normal adults [49]. Another longitudinal study also found that changes in tau were more strongly linked to the rate of change in Aβ deposition levels. They also noted that tau changes occurred soon after Aβ positivity was detected [50]. Their findings demonstrated that the association between Aβ deposition and cognition was delayed and indirect and was mediated by tau. Additionally, individuals with a low change rate in both Aβ and tau showed stable cognition. Widespread tau in HC subjects and acceleration phase through distal association with Aβ critically important for a better understanding of tau spreading, especially development in disease-related brain regions. Surprisingly, the MCI/mild AD subject’s tau in the acceleration phase shows a weak distal association with Aβ. These weak associations could be explained by the fact that the rate of deposition of Aβ in disease levels decreases at a higher accumulation level [51, 52].

Lastly, we found a weak distal association between tau and Aβ in the post-acceleration phase of participants, which included more than 96% MCI or mild AD and Aβ+ subjects. While the level of deposition and the spatial distribution of tau and Aβ strongly increase in this phase compared with the acceleration phase, no distal association survived after multiple comparison corrections. Earlier studies have reported that the rate of Aβ accumulation seems to decelerate as it reaches higher levels [51, 52]. Previous neuropathological studies also demonstrated that deposition of tau and Aβ rate decrease with aging in AD patients [53]. Furthermore, diverse studies reported a significant negative age association with tau deposition in the later stage of accumulation at the disease level [53, 54]. The rate change in the deposition in elderly patients with dementia would explain the results of MCI or mild AD subjects in the acceleration and post-acceleration phases. The alternative explanation is the difference between the pathological basis between the early and late stages of the disease [53, 55]. The hypothesis we want to highlight is based on the neural connection between different regions in the brain, especially in the distal association. The target regions like the entorhinal cortex that shows the distal association with Aβ in other cortical regions in acceleration phase subjects are functionally connected [56]. On the other hand, tau and Aβ both affect the functional connectivity [57, 58], and these two pathologies might induce a disruption in the functional connectivity in late stages of deposition and finally lead to an uncoupled with one another.

The results of this study emphasize the strong distal association between the two key proteinopathies of AD. While the cellular mechanism of this association is still elusive, several in vitro and in vivo studies have demonstrated that Aβ triggers the tau deposition [59-63]. It has been shown that Aβ enhanced tau deposition not only at the site of injection but also in distal and functionally connected brain regions to the injection site [64]. The valid mechanism for explaining the association between tau and Aβ deposition is the inducement of neural connections. Previous studies indicated that neural connection hubs depend on the tau based on their weighted degree or connectivity [42, 44], which means that hyperactivity accelerates the progression of pathological tau along vulnerable neural connections. On the other hand, several reports indicated the influence of Aβ deposition on hyperactivation on the neural connection in mouse models [65-68]. Furthermore, it has been shown in MCI patients that the Aβ deposition leads to hyperactivation in the hippocampus [69, 70]. By this evidence, we can suggest the mechanism for the distal association between tau and Aβ that is induced by neural connections: Aβ deposition is associated with a disruption in neural connection that causes hyperactivity; this hyperactivation then enhances the tau deposition distally throughout the brain regions that functionally connected with each other; in disease stage, this neural connection gets disrupted and leads to weak associations. Notably, PET can only detect fibrillar aggregations of Aβ; soluble or intracellular aggregations could also play a critical role in the distal association. Since the utility of anti-amyloid drugs in the later stages of AD remains unclear, understanding the early effects of Aβ on progressive tau deposition in the brain is critical for preventing AD progression. Thus, a longitudinal study is essential for understanding the underlying pathophysiology explaining the association between these two pathologies.

There are important limitations and areas for further investigation from the present study that need to be discussed. The first limitation of this study is that a different Aβ PET tracer was used for healthy controls (18F-Florbetaben) and individuals with MCI/mild AD (18F-Florbetapir). To address this issue, all analyses in the study were replicated using centiloid standard values instead of Aβ SUVR [71]. Furthermore, we also implemented interclass statistical analysis to compare the centiloid and SUVR regional values. The strong average correlation of 0.99 and the standard deviation of 0.01 was calculated across 68 regions, and none of the reported results changed by centiloid values. Since the tau PET tracer was the same in all subjects and several results were based on only HC or only MCI/mild AD subjects separately, in this paper, only analyses using SUVR measures were reported for both pathologies to ensure greater comparability and ease of understanding. Next, the relationship between AD pathologies is not easy to analyze in a cross-sectional and group-wise manner. Thus, considering the pathological changes individually in a longitudinal dataset is necessary. Finally, while 572 older and 144 younger samples were used in this study, greater than typically reported in human studies in the field, especially with the second generation of PET tracers, it is possible that our sample size was not large enough to provide sufficient statistical power to detect all associations in our regression analyses.

### Conclusion

The results of the current study illustrate the robust distal association between tau and Aβ, not only in the late stage (acceleration phase) but also strongly in the early stages of tau deposition (pre-acceleration phase), particularly in the entorhinal cortex and parahippocampal gyrus regions. Our study shows that the distal association begins with MTL tau during the early stage and is evident in the later stages of tau accumulation. It is interesting to note that distal association was attenuated in symptomatic subjects with cognitive decline (MCI and mild AD subjects), whereas this association is strongly enhanced in Aβ+ asymptomatic subjects (HC). Since the failure of anti-amyloid drugs may be linked to delayed initiation of treatments, understanding distal association in the early and late stages of the disease is crucial to improving drug development and preventing AD progression.

## Data Availability

All data produced in the present study are available upon reasonable request to the authors.

